# Serial interval, basic reproduction number and prediction of COVID-19 epidemic size in Jodhpur, India

**DOI:** 10.1101/2020.07.03.20146167

**Authors:** Suman Saurabh, Mahendra Kumar Verma, Vaishali Gautam, Akhil Goel, Manoj Kumar Gupta, Pankaj Bhardwaj, Sanjeev Misra

## Abstract

**Background:** Understanding the epidemiology of COVID-19 is important for design of effective control measures at local level. We aimed to estimate the serial interval and basic reproduction number for Jodhpur, India and to use it for prediction of epidemic size for next one month.

**Methods:** Contact tracing of SARS-CoV-2 infected individuals was done to obtain the serial intervals. Aggregate and instantaneous *R*0 values were derived and epidemic projection was done using R software v4.0.0.

**Results:** From among 79 infector-infectee pairs, the estimated median and 95 percentile values of serial interval were 5.98 days (95% CI 5.39 – 6.65) and 13.17 days (95% CI 11.27 – 15.57), respectively. The overall *R*0 value in the first 30 days of outbreak was 1.64 (95% CI 1.12 – 2.25) which subsequently decreased to 1.07 (95% CI 1.06 – 1.09). The instantaneous *R*0 value over 14 days window ranged from a peak of 3.71 (95% CI 1.85 -2.08) to 0.88 (95% CI 0.81 – 0.96) as on 24 June 2020. The projected COVID-19 case-load over next one month was 1881 individuals. Reduction of *R*0 from 1.17 to 1.085 could result in 23% reduction in projected epidemic size over the next one month.

**Conclusion:** Aggressive testing, contact-tracing and isolation of infected individuals in Jodhpur district resulted in reduction of *R*0. Further strengthening of control measures could lead to substantial reduction of COVID-19 epidemic size. A data-driven strategy was found useful in surge capacity planning and guiding the public health strategy at local level.

## Introduction

COVID-19 has emerged as the largest pandemic of 21^st^ century with 10.5 million confirmed cases and half million deaths worldwide, as on 2 July 2020.^1^ India has become the fourth most affected country worldwide with around 0.6 million confirmed COVID-19 cases.^1^ COVID-19 is an emerging infectious disease with the onset of symptoms of first case having been reported from Wuhan, China in December 2019.^2^ Various epidemiological studies are being done to understand the transmission dynamics of the disease. Consequently, the estimated parameters such as serial interval and basic reproductive number (R0) are being used to guide the control strategies and to enable disease forecasting.^3–5^

In the early phase of the COVID-19 pandemic, India had adopted the policy of universal health-facility based isolation of all SARS-CoV-2 infected individuals irrespective of symptomatic status. However, in view of the increasing number of COVID-19 cases, home isolation of asymptomatic and mild cases was introduced on 10 May 2020.^6^ Therefore, it is important to achieve an epidemiological understanding of COVID-19 situation at district level in the changed scenario so that it could be used to guide control measures and surge preparedness on a real-time basis. Jodhpur is situated in the western part of India in Rajasthan state (Fig 1). The first COVID-19 case was an imported case reported on 9 March 2020 in Jodhpur, India.

**Fig 1:**
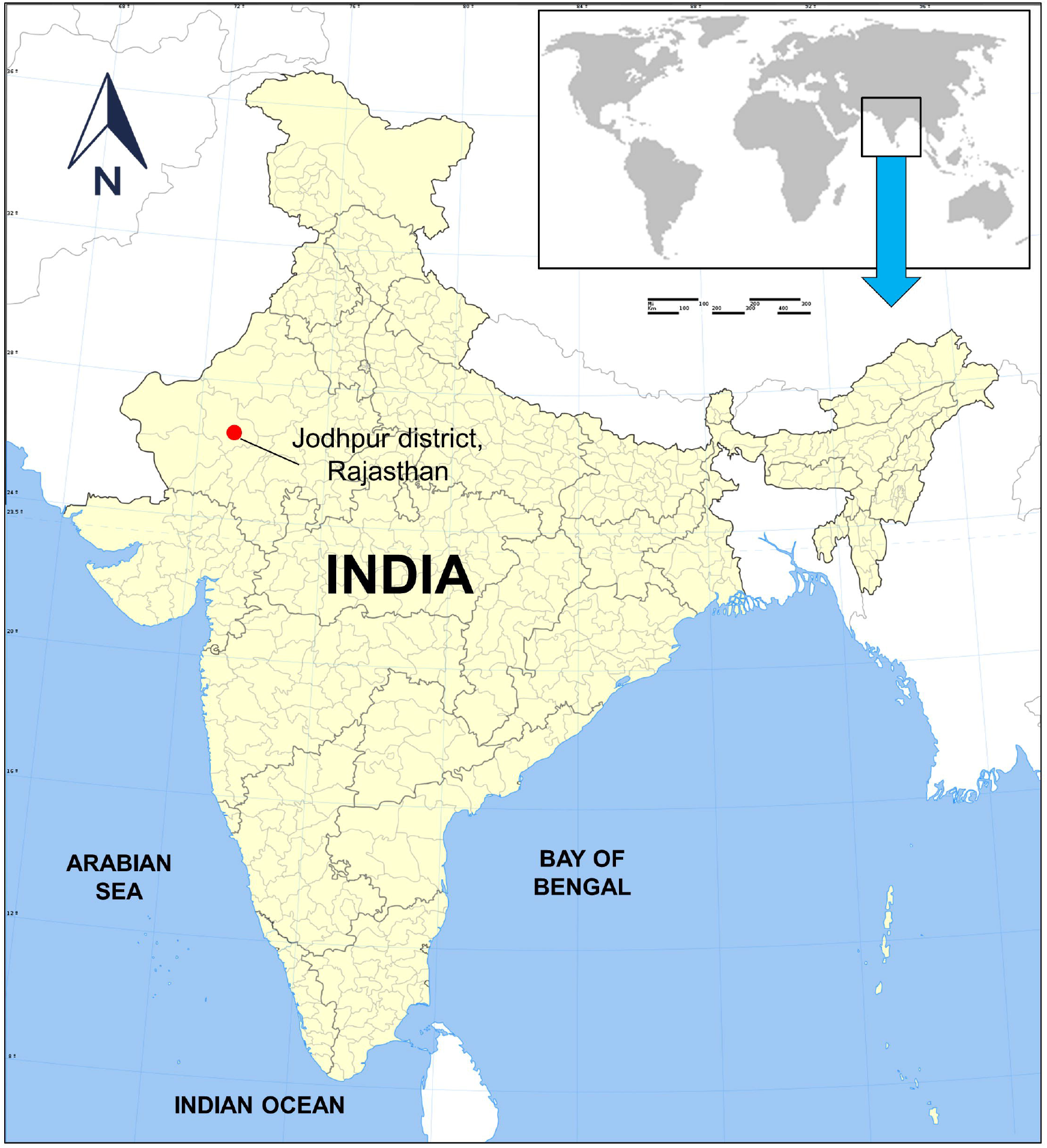
Location of Jodhpur district within Rajasthan state of India *(Modified from source file - https://commons.wikimedia.org/wiki/File:India_districts_map.svg, Creative Commons Attribution-Share Alike 4.0 International license)*

## Methods

### Serial interval estimation

Individuals meeting suspect case definition for COVID-19 were tested with rRT-PCR (Real-Time Reverse Transcription-Polymerase Chain Reaction) at our institute in Jodhpur, India as per the national guidelines.^7^ Those found positive for SARS-CoV-2, were further assessed for their contact history with known COVID-19 cases in their household. Serial interval was estimated based on the time duration between the symptom onsets of the infector-infectee pairs thus identified. For asymptomatic individuals, the date of collection of first positive sample was taken as a proxy for symptom onset. Mean and standard deviation of serial interval was calculated.

Further, the serial interval data was fitted to weibull, log-normal, log-logistic and generalized gamma distributions using Flexsurv package in R software version 4.0.0. ^8^ The estimates of median serial interval were taken from the best fitting model based on minimum Akaike Information Criterion (AIC) value. Standard maximum likelihood approach was used to obtain the best model fit to actual data.

### Estimation of R0

The basic reproduction number (*R*0) is defined as the average number of susceptible individuals infected by a single primary case.^9^ The daily COVID-19 case data of Jodhpur district was converted to incidence object using Incidence package in R software.^10^ EarlyR and EpiEstim packages in R software were used to estimate overall and instantaneous values of basic reproduction number using the parameter estimates of serial interval, respectively.^11,12^ Instantaneous *R*0 values were calculated based on method of estimating daily incidence based on a Poisson process determined by daily infectiousness, as proposed by Jombart and Nouvellet *et al*.^10,13^ Here *λ*_*t*_, the force of infection observed on day t is expressed by the following equation:

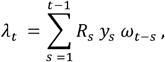

where *y*_*s*_ is the incidence of cases on day *s. R*_*s*_ is the instantaneous reproduction number on day *s*. The value of *ω*_*t-s*_ is the probability mass distribution of the serial interval which represents the infectiousness of incident cases on day *s* in order to result in secondary cases on day *t*. As a practical approach used by earlier studies, we approximated day of reporting of the case as the day of onset, in the absence of exhaustive symptomatic history of each reported case.^13^

We also used another method by Wallinga and Teunis for estimation of the time varying *R*0 based on probability of transmission between infector-infectee pairs.^14^ We used the parametric method of specifying the mean and standard deviation of serial interval distribution for both the methods. Time window of both 7 days and 14 days was used for calculation of instantaneous *R*0.

### Forecasting of the epidemic size

Forecasting of daily and cumulative COVID-19 cases for the next 30 days was done based on the overall *R*0 value and based on *R*0 value of the past 30 days as input parameters using the Projections package in R.^10^ As required, serial interval distribution was specified as scale and shape parameters of gamma distribution.

Forecasting of daily incidence was based on a Poisson process determined by daily infectiousness.^13^ The specified serial interval distribution is taken as a prior while utilizing the Bayesian methodology for Markov Chain Monte Carlo (MCMC) sampling using the Metropolis algorithm. The 95% confidence intervals of projected daily and cumulative incidence were calculated using bootstrap resampling method with 1000 samples.

## Results

### Serial interval

Till 24 June 2020, 2619 cases were reported from the district (Fig 2). Contacts of 522 SARS-CoV-2 infected individuals were traced from 15 April – 20 June 2020 and among them 91 individuals had a positive contact history with a confirmed COVID-19 case. Among them, serial interval data for 79 infector-infectee pairs was obtained (supplementary table 1). The mean serial interval was 6.75 days with a standard deviation of 3.76 days. The log-normal distribution was found to be the best fitting with serial interval with minimum Akaike Information Criterion value (Fig 3). The median and 95 percentile values of serial interval were 5.98 days (95% CI 5.39 – 6.65) and 13.17 days (95% CI 11.27 – 15.57), respectively estimated from the fitted log-normal distribution.

**Fig 2:**
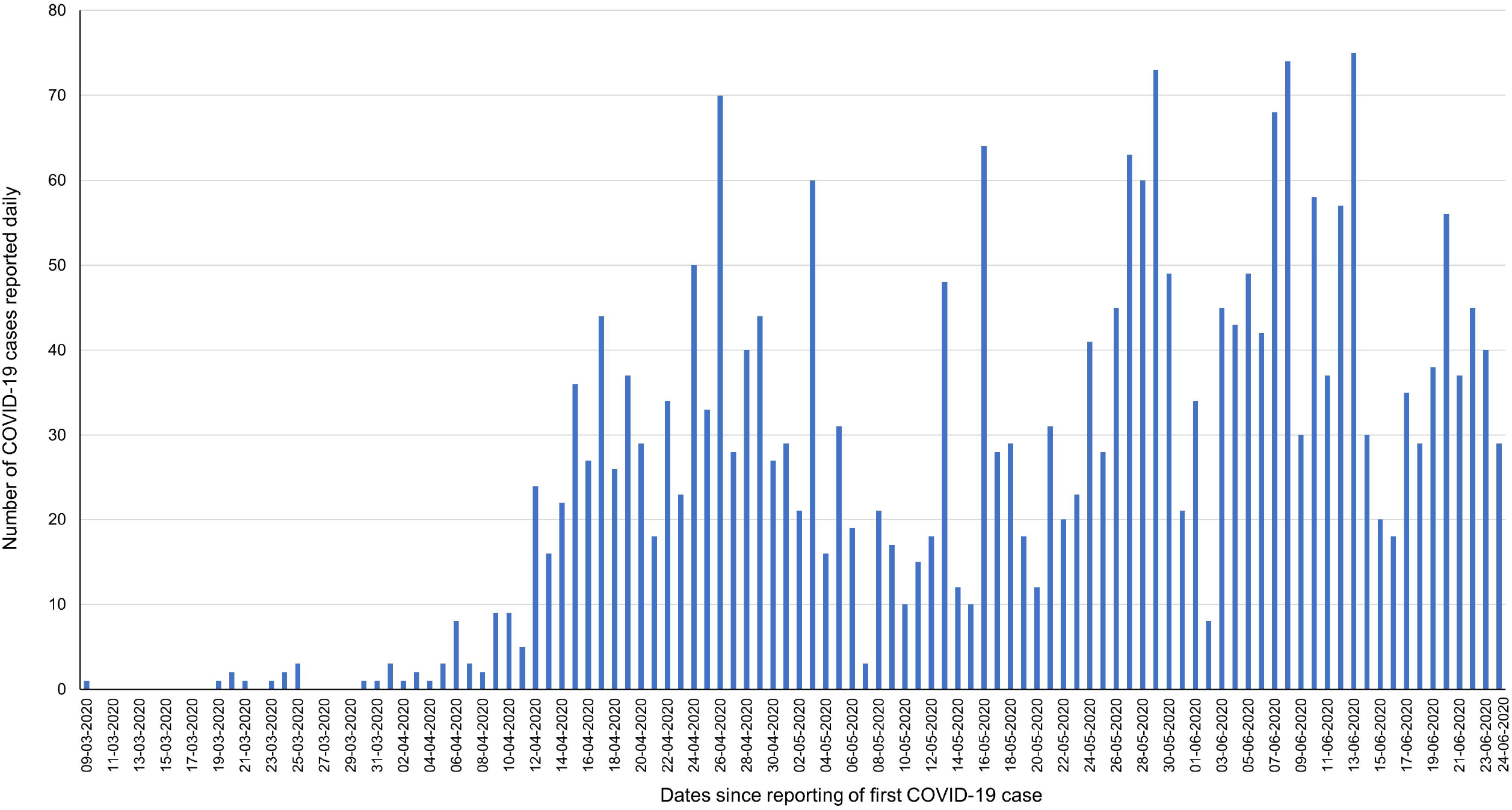
Daily COVID-19 cases at Jodhpur, India from 9 March 2020- 24 June 2020

**Fig 3:**
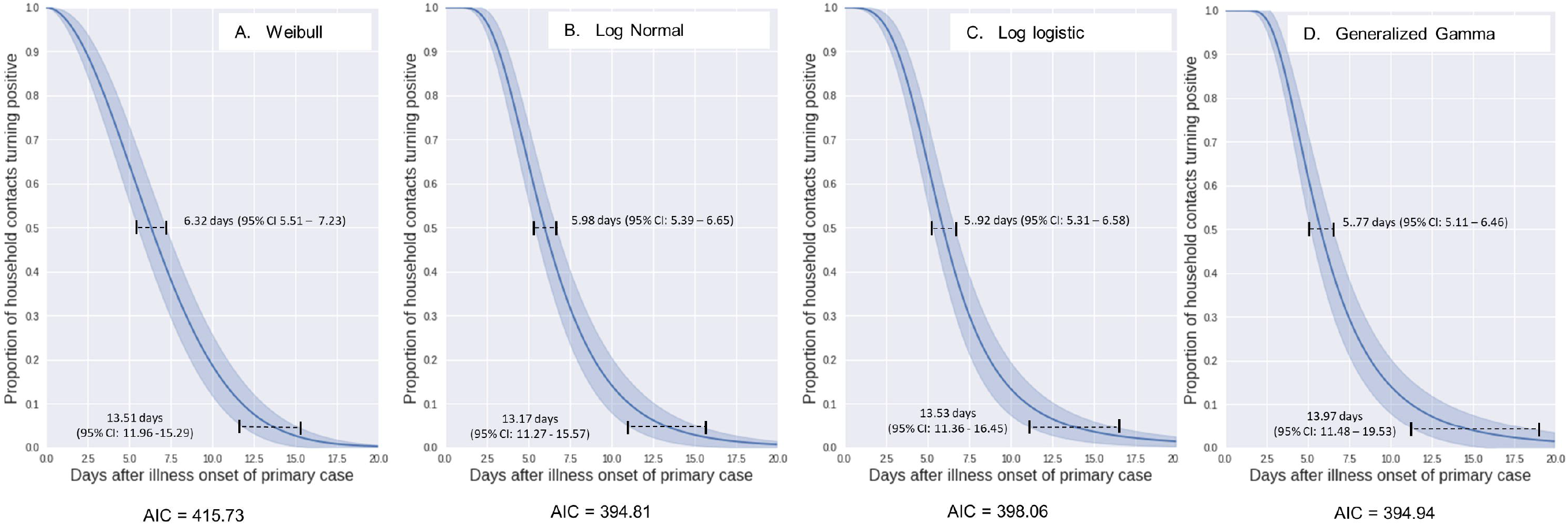
Estimates of median and 95 percentile of serial interval data as fitted to weibull, log-normal, log-logistic and generalized gamma distributions (n = 79 pairs)

### Estimation of R0

The overall *R*0 value in the first 30 days after reporting of first case was 1.64 (95% CI 1.12 – 2.25) which subsequently decreased to 1.07 (95% CI 1.06 – 1.09). The overall R0 value for the entire duration of 9 March – 24 June 2020 was 1.07 (95% CI 1.06 – 1.09), whereas it was 1.17 (95% CI 1.06 -1.23) for the last 30 days.

The instantaneous *R*0 value calculated using the method by Jombart and Nouvellet *et al* yielded maximum values of 6.48 (95% CI 2.10 – 13.27) and 3.71 (95% CI 1.85 - 2.08) using sliding time-windows of 7 days and 14 days respectively (Fig 4).

**Fig 4:**
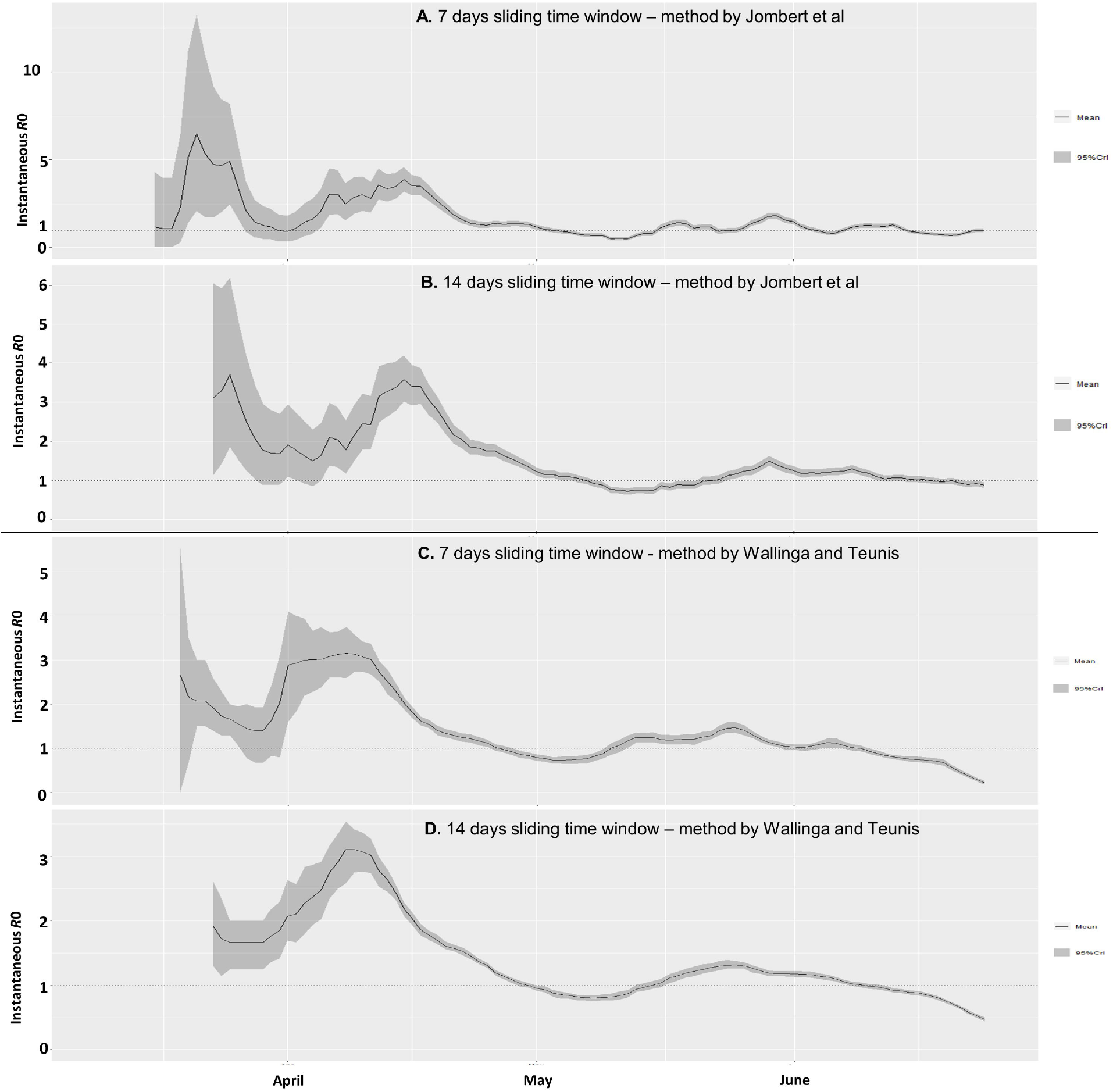
Instantaneous R0 values for Jodhpur, India estimated using method by Jombart et al (a-b) and by Wallinga and Teunis (c-d) using time windows of 7 and 14 days

Similarly, using the method developed by Wallinga and Teunis the maximum values of instantaneous R0 were 3.16 (95% CI 2.60 – 3.75) and 3.11 (95% CI 2.75 – 3.41), respectively (Fig 4). The latest instantaneous *R*0 value estimated on 24 June 2020, using the method by Jombart and Nouvellet *et al* were 0.99 (95% CI 0.88 – 1.11) and 0.88 (95% CI 0.81 – 0.96) taking 7- and 14-days sliding time-windows, respectively. Similarly, the latest instantaneous *R*0 values estimated on 24 June 2020, using the method by Walling and Teunis were 0.22 (95% CI 0.18 – 0.27) and 0.47 (95% CI 0.44 – 0.50) taking 7- and 14-days sliding time-windows, respectively.

### Projection of epidemic size

The number daily cases projected for the next month while taking a *R*0 value of 1.07 (representing the entire duration of transmission from 9 March – 24 June 2020) ranged from 44 individuals (95% CI 32 -55) on day 1 to 64 individuals (95% CI 42 – 87) on day 30 (Fig 5). Similarly, the number daily cases projected for the next month while taking a *R*0 value of 1.17 (representing only the last 30 days prior to 24 June 2020) ranged from 46 individuals (95% CI 34 - 59) on day 1 to 85 individuals (95% CI 60 – 111) on day 30 (Fig 5). The cumulative projection of number of COVID-19 case over the next 30 days while taking *R*0 value of 1.07 was 1563 individuals (95% CI 1281 – 1845). Similarly, the projection over next one month was 1881 individuals (95% CI 1542 – 2220) with input of *R*0 value of 1.17 considering the transmission most recent 30-days period. A scenario of 50% reduction in transmissibility above the maintenance level of *R*0 (i.e. from *R*0 of 1.17 to 1.085) assuming further strengthening of control measures resulted in 1450 (95% CI 1151 – 1750) cumulative cases over the next month, corresponding to 23% reduction in projected case-load.

**Fig 5:**
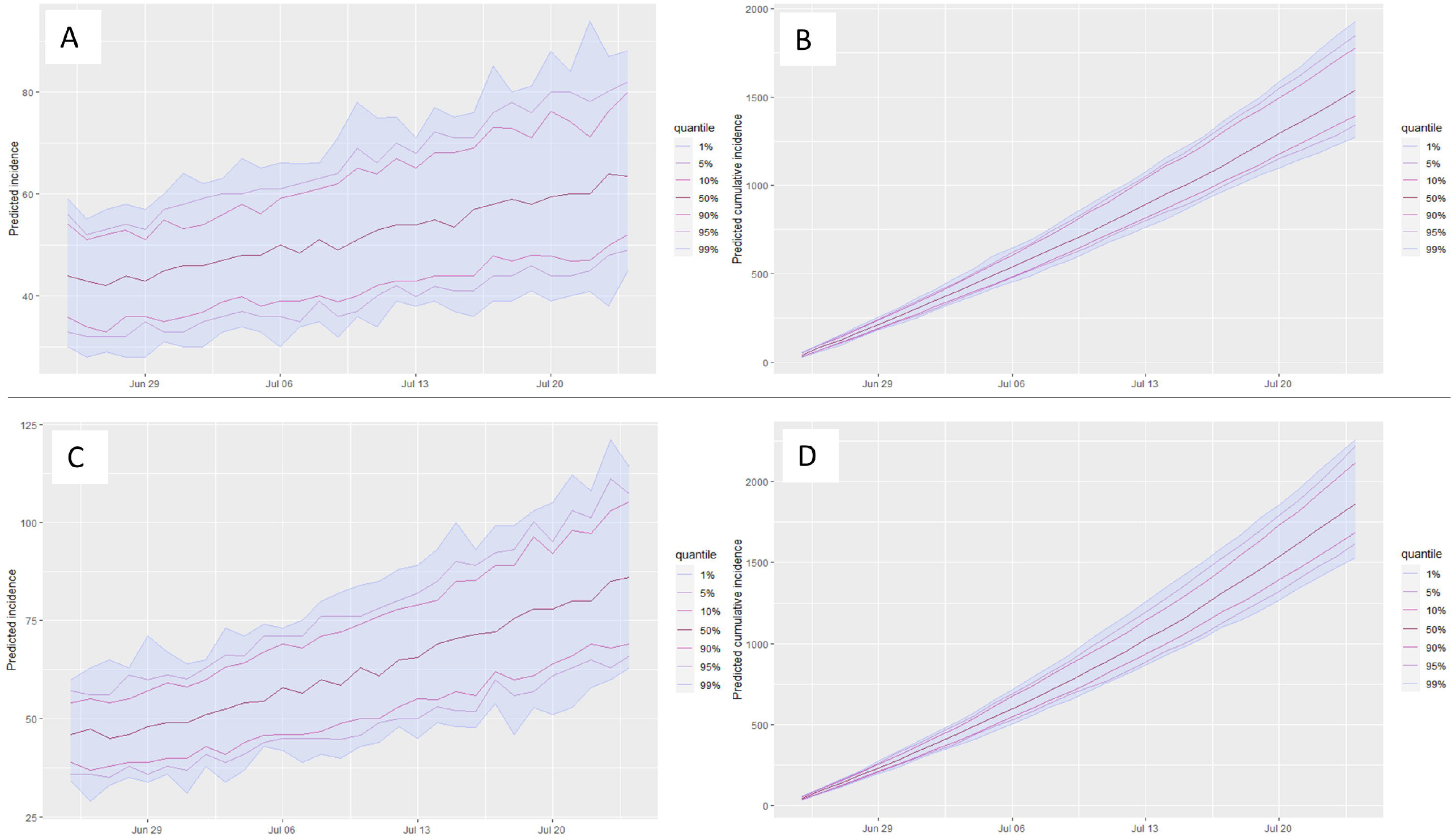
Projection of daily and cumulative COVID-19 case-load over the next 30 days using R0 value of 1.07 for overall duration till 24 June 2020 (a - b) and R0 value of 1.17 for one month preceding 24 June 2020 (c - d)

## Discussion

### Implications of serial interval and R0 estimation

Our observation of mean serial interval fell within the range of 4-8 days estimated by a meta-analysis of 7 studies conducted during the early phase of the COVID-19 pandemic.^15^ Another meta-analysis including studies only from China estimated a range of serial interval from 4.10 – 7.5 days.^16^ Our experience suggests that the median and 95% confidence interval estimate of serial interval should be reported alongside the mean and standard deviation as the former approach is more susceptible to be influenced by extreme values. It has also been suggested that longer serial interval intervals can be noted due to preventive interventions and during the course of the epidemic.^17,18^ Therefore, it is preferable to estimate recent serial interval locally to better understand the transmission of SARS-CoV-2.

The distribution of *R*0 values was consistent with the observation from other countries indicating a similar transmission pattern.^4,18^ Once the peak of *R*0 value was reached in the first week of April, subsequent reduction towards April end could be attributed to aggressive testing, contact tracing and isolation measures in the urban area of Jodhpur during the April month. Earlier detection of infection followed by isolation is known to reduce the *R*0 value through limiting both the duration of effective contact and the number of susceptibles an infected individual can come in contact with.^9^ Our findings further support that parameters such as serial interval, incubation period and *R*0 values are likely to vary throughout the course of the epidemic and will depend on the local factors influencing transmission such as demographics, environmental conditions, modelling methodology and the stringency of the control measures.^9,19^

### Projection of epidemic

The projected estimate of daily case and the final outbreak size were found to depend on the value of *R*0 entered in the model.^20,21^ The method used to estimate the *R*0 value and the time-window over which *R*0 was calculated influenced the final projection by a wide margin. The 14-days-time window yielded less variable instantaneous *R*0 estimates as compared to a 7-days-time window. We found that the method by Wallinga and Teunis was more sensitive to recent fluctuations in daily case count, as compared to the method by Jombart *et al*, while taking the same time window. Further, as per the renewal equation stated earlier, the values of *R*0 are most influenced by the trend in daily cases reported within the range of the serial interval i.e. within 5-6 days. This also pre-assumes homogenous mixing, which becomes less applicable with larger populations with cases emerging from widely separated clusters. Also, the impact of methods of *R*0 estimation and time windows were more pronounced when there was a fluctuating trend in cases or the *R*0 value was close to 1. Therefore, we recommend that *R*0 values over a comparatively larger period be taken instead of instantaneous *R*0 values for providing reliable projections in larger populations.

### Strengths and limitations

One of the strengths of our study was estimation of serial interval based on large data over a period of two months. Also, since our study was based on contact history of infected individuals instead of daily follow-up of contacts of infected individuals for disease onset, we minimized underreporting of longer serial intervals which is possible due to right-truncation in assessing serial interval based on follow-up method.^22^ Further, our use of time-varying method for daily *R*0 estimation and maximum likelihood method for overall *R*0 estimation had the benefit of less bias as compared to exponential growth and sequential Bayesian methods.^23^ The time-varying method had the added advantage of providing daily *R*0 values which were useful in assessing the effectiveness of control measures, as compared to other methods which provide only an aggregate *R*0 value.^22^

Population level estimates relying on daily reports could underestimate the value of *R*0 as compared to those of closed populations, as many infected individuals are likely to be missed, especially if the testing capacity is limited or proportion of asymptomatics is high.^20^ Further, modelling assumptions such as assuming a finite probability of interaction of infector-infectee pairs reported within a range of serial interval might not be applicable for large population cohorts.^14^ In order to overcome such limitations use of both spatial and temporally structured data has been proposed.^24^

## Conclusions

Public health measures such as testing, contact tracing and home isolation were found to reduce to instantaneous *R*0 value and could thereby reduce the final outbreak size. The final epidemic size was found to be influenced by *R*0 values, which in turn depended on the stringency of control measures. Even a marginal reduction in *R*0 as a result of strengthening control measures was found to considerably reduce the projected COVID-19 burden in Jodhpur, India. Projections are feasible based on publicly released daily COVID-19 case data and could be useful in guiding a data-driven COVID-19 response strategy at a local level. This could be utilized for both surge capacity planning of number of hospital beds and ventilators required, and also for the public health response such as number of staff required for contact tracing and for provisioning of institutional quarantine or isolation facility. Therefore, considering the increasing case load and dynamic situation of COVID-19, a decentralized evidence-driven approach appears to be the need of the hour.

## Data Availability

Data on which the study is based is included in the manuscript and in the supplementary files.

## AUTHOR STATEMENTS

### Authors’ contributions

MKV, VG and SS collected the data and SS conducted the analysis. SS wrote the draft manuscript with further inputs from MKV, VG, AK, MKG and PB. PB coordinated the data collection process. SM provided overall supervision of the lab testing, clinical care and research related to COVID-19 at AIIMS - Jodhpur, India. All authors approved the final manuscript.

## Acknowledgements

We acknowledge the district administration Jodhpur for providing the daily COVID-19 case data.

## Ethical approval

The study has been approved by the Institutional Ethics Committee of All India Institute of Medical Sciences (AIIMS) - Jodhpur, India.

## Supplementary file

**Supplementary table 1: Data for ‘Serial interval estimation of COVID-19 in Jodhpur, India’**

